# AGG Repeat Expansion and Aggregation of BIN1 in Multiple System Atrophy

**DOI:** 10.1101/2025.09.02.25334695

**Authors:** Kodai Kume, Takashi Kurashige, Takeshi Itabashi, Akio Akagi, Takashi Ando, Masataka Nakamura, Mai Kikumoto, Atsushi Tamada, Masaki Kamada, Takashi Ayaki, Yuishin Izumi, Ichiro Yabe, Keiko Muguruma, Tatsuo Miyamoto, Yasuhi Iwasaki, Hideshi Kawakami

## Abstract

Multiple system atrophy (MSA) is a fatal, sporadic α-synucleinopathy characterized by glial cytoplasmic inclusions (GCIs) in oligodendrocytes. No causative gene for MSA has been identified to date. Herein, we report an intronic AGG repeat expansion in *BIN1*, a major susceptibility gene for Alzheimer’s disease, in a familial MSA case and 13% of autopsy-confirmed sporadic cases. In one case, the expansion was detected only in the cerebellum, implicating somatic mosaicism in disease pathogenesis. The BIN1 protein aggregate localized to GCIs and associated with α-synuclein fibrils. Notably, BIN1 was absent from Lewy bodies, highlighting a potential MSA-specific role. Our findings indicate a novel somatically unstable repeat expansion contributing to MSA and support the notion that BIN1 aggregation plays a central role in disease-specific protein pathology.

**One-Sentence Summary:** Repeat expansion and accumulation of BIN1 play an important role in the pathogenesis of multiple system atrophy.

## Main Text

Multiple system atrophy (MSA) is a neurodegenerative disorder characterized by autonomic failure, parkinsonism, and cerebellar ataxia^1^. Pathologically, MSA is defined by glial cytoplasmic inclusions (GCIs)^2,3^ that accumulate phosphorylated α-synuclein^4^. Although most cases are sporadic, variants in *SNCA*^5^, *LRRK2*^6^, and *COQ2*^7^, together with genome-wide association signals at *ZIC4*^8^, *USP38-DT, KCTD7*, and *lnc-KCTD7-2*^9^ modulate risk, leaving the primary genetic architecture of MSA unresolved and the molecular basis of GCI formation obscure.

In this study, we demonstrate that expanded AGG trinucleotide repeats in intron 1 of *BIN1* act as a causative variant of MSA and that BIN1 protein is an obligate component of GCIs irrespective of repeat status. The findings of this study establish *BIN1* repeat expansion as a genetic driver of MSA and highlight BIN1 misfolding as a convergent pathogenic mechanism, providing a direct molecular link between oligodendroglial inclusion and neurodegeneration.

## Results

### Genetic analysis of familial multiple system atrophy

We analyzed a patient with MSA (II-2) whose younger sibling (II-3) died of MSA in his 60s (Fig. 1A). The patient developed gait instability in her 70s, with progressive worsening of mobility. She experienced upper limb incoordination and dysarthria, and was diagnosed with MSA-cerebellar type (MSA-C) at a neurology clinic. She also reported episodes of lightheadedness and vocalizations during sleep. Brain MRI scans of both siblings showed the characteristic “hot cross bun” sign and atrophy of the pons and cerebellum, supporting the diagnosis of MSA (Fig. 1B).

**Fig. 1.**
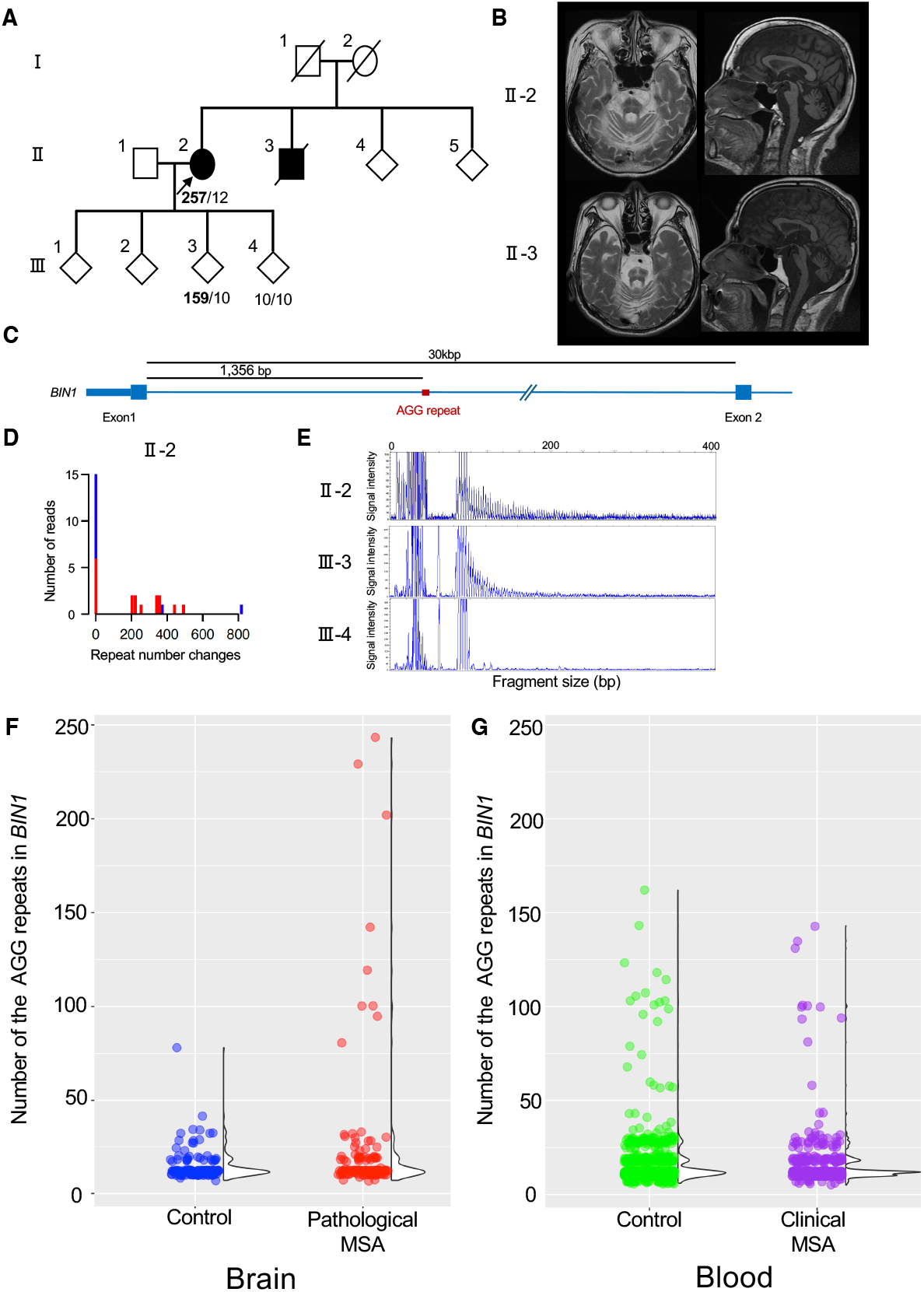
MSA family and AGG repeat expansion in *BIN1*. **A**, Pedigree of the MSA family. The arrow indicates the index patient. Numerical values represent the number of AGG repeats in *BIN1*, with the expansion allele shown in bold. **B**, Brain MRI scan of two affected family members. The left panels show the axial view of the T2-weighted image, whereas the right panel displays a sagittal view of the T1-weighted image. **C**, Diagram illustrating the AGG repeat expansion in intron 1 of *BIN1*. **D**, Histogram of repeat number changes analyzed by long-read sequencing for individual II-2. Red bars represent sequencing reads of the positive strand, and blue bars represent the negative strand. **E**, Fragment analysis of repeat-primed PCR for AGG repeats in *BIN1*. Individuals II-2 and II-3 display sawtooth patterns. **F, G**, The distribution of repeat number in *BIN1* for brain control (N = 65), pathological MSA (N = 67), blood control (N = 574), and clinical MSA (N = 224). The value denotes the repeat number in a single allele. Half-split violin plots display the probability density of the data.

We first performed whole-exome sequencing on individual II-2 and confirmed the absence of variants in *the SNCA*^5^, *LRRK2*^6^, and *COQ2*^7^genes, which are previously reported to be associated with MSA. To assess potential repeat expansions, we conducted whole-genome sequencing using a PromethION long-read sequencer. Analysis with tandem-genotypes software^10^ identified an expansion of the AGG repeat in intron 1 of *BIN1* (chr2:127,105,468-127,105,503 [hg38], Fig. 1C). *BIN1* is a causative gene for centronuclear myopathy^11^ and the second most associated locus for Alzheimer’s disease, and is mainly expressed in oligodendrocytes in the brain^12,13^. BIN1 possesses the N-terminal BIN1/Amphiphysin/Rvs domain and a C-terminal MYC-Binding Domain, with functions related to endocytosis, presynaptic vesicle release dynamics, and the inflammatory response in the central nervous system^13^. The AGG repeat length ranged from 200 to 800 repeats, suggesting an unstable repeat (Fig. 1D). Utilizing long-read sequencing data, we constructed a consensus sequence of the repeat, indicating a representative repeat length of 257 repeats (Supplementary Fig. S1). We also assessed *BIN1* repeat expansions in other family members (III-3 and III-4) via repeat-primed PCR. Like individual II-2, individual III-3 exhibited a saw-tooth pattern in the assay, suggesting a repeat expansion (Fig. 1E). Cas9-mediated enrichment sequencing confirmed that the repeat length from III-3 was 159 repeats. During germline transmission, the repeat length decreased from 257 to 159 repeats (Fig. 1A), demonstrating the instability of the AGG repeats in *BIN1*. Individual III-3 (age 50s) exhibited no neurological symptoms, but continued observation is required to monitor potential MSA development.

### Screening of the repeat expansion for MSA

Of 67 patients with pathological MSA, 9 (13.4%) carried repeat expansions exceeding 80 repeats as compared to none (0%) of 65 brain controls (odds ratio, infinity; 95% confidence interval [CI], 2.1 to infinity; *p* = 0.003 by Fisher’s exact test) (Fig. 1F and Supplementary Table S1). Among the 224 patients with clinical MSA, 10 (4.5%) carried the repeat expansions as compared with 14 of 574 blood controls (odds ratio, 1.87; 95% CI, 0.8 to 4.5; *p* = 0.16 by Fisher’s exact test) (Fig. 1G and Supplementary Table S1). The AGG repeat expansion in *BIN1* was identified in the MSA-parkinsonian type (MSA-P) and the MSA-cerebellar type (MSA-C).

### MSA with somatic AGG repeat expansion in *BIN1*

We investigated the repeat expansions in *BIN1* in six additional patients with MSA from another cohort, identifying one patient with a positive repeat-primed PCR result. An expanded repeat (91 repeats) was identified only in the cerebellum, indicating that the repeat expansions occurred as a somatic mutation in this patient (Fig. 2). The peaks of slightly expanded repeats were observed in the cervical spinal cord and frontal lobe (Fig. 2), suggesting that the repeat expansion in *BIN1* is unstable in the central nervous system.

**Fig. 2.**
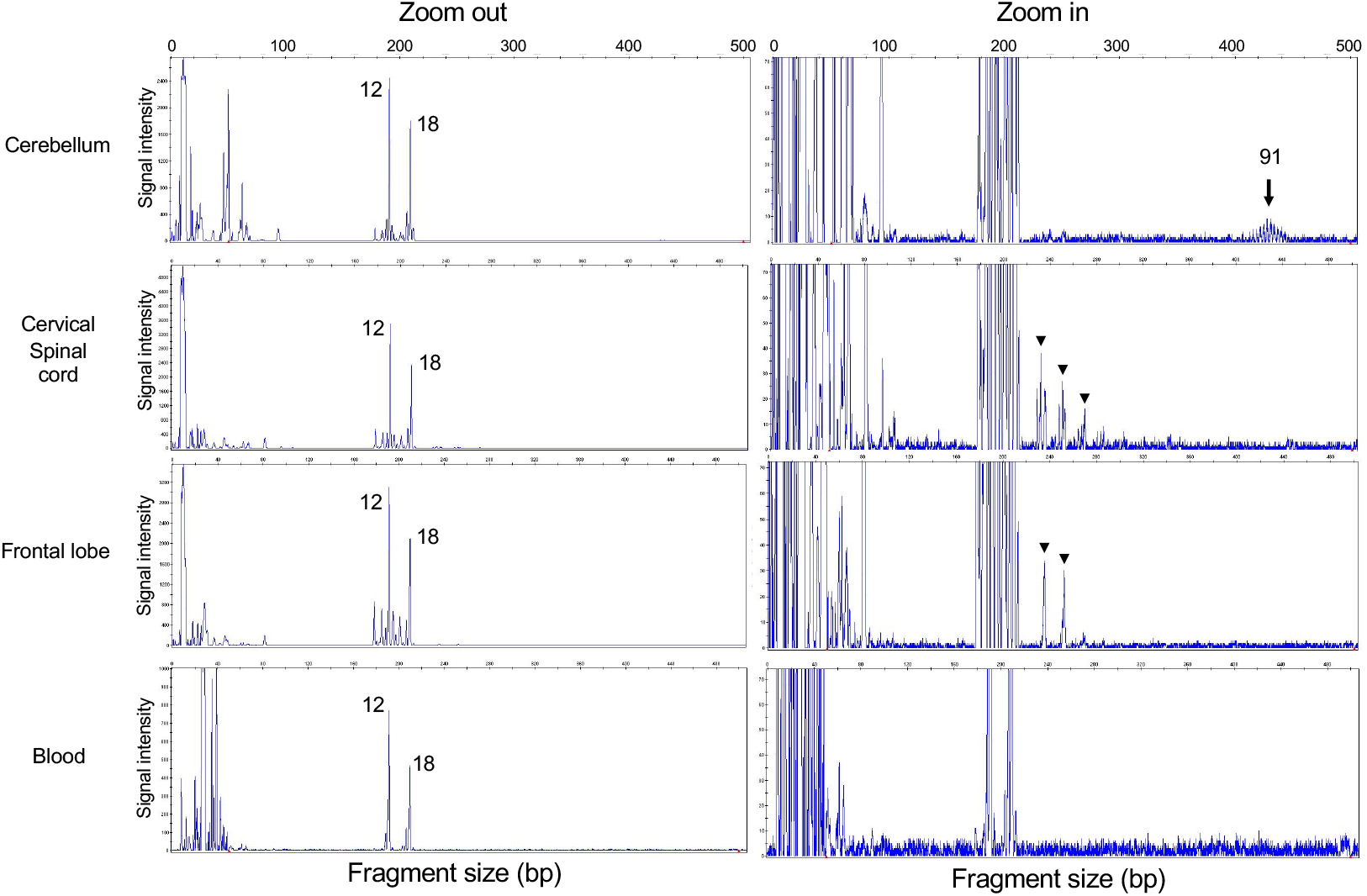
MSA with somatic repeat expansion in *BIN1*. Fragment analysis of flanking PCR for AGG repeat in *BIN1* using DNA from the cerebellum, cervical spinal cord, frontal lobe, and blood. Numerical values indicate the number of repeats. The arrow highlights the signal corresponding to the somatic repeat expansion, whereas the arrowheads point to peaks representing slightly expanded repeats.

Subsequent histopathological examinations of the cerebellum, pons, and frontal cortex in the patient revealed BIN1-positive GCIs exclusively in the cerebellum, with occasional inclusions in the frontal cortex. No neuronal inclusions were observed in any of the areas examined (Supplementary Fig. S2).

### Histopathological manifestations of MSA with AGG repeat expansion in *BIN1*

A summary of the patients with MSA is provided in Supplementary Table S2. Initially, we observed myelin pallor in the cerebellar white matter lesions of patients with MSA (R+) using Klüver–Barrera (KB) staining (Fig. 3A). This finding was not present in patients with MSA (R−), except in those with complicating cerebellar infarcts. However, Gallyas–Braak silver (GB) staining did not show differences in the presence of GCIs in the cerebellar white matter lesions or neuronal cytoplasmic inclusions (NCIs) in the dentate nuclei or Purkinje cells between MSA (R+) and MSA (R−) (Supplementary Fig. S3A, B).

**Fig. 3.**
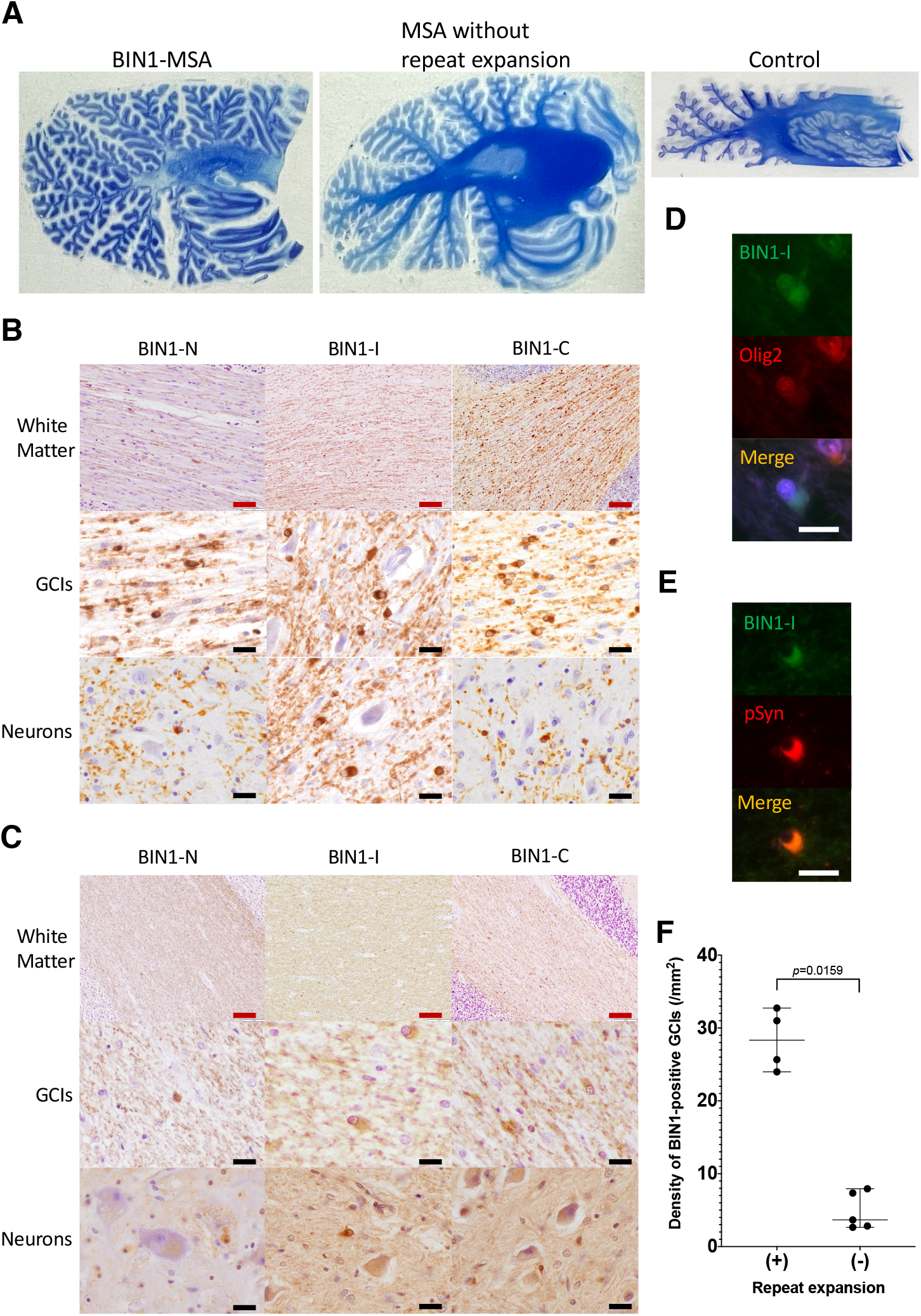
Pathological manifestation of MSA with AGG repeat expansions in *BIN1*. **A**, Kruver–Barrera staining of MSA (R+), MSA (R−), and control tissue. Myelin pallor in white matter lesions was observed in MSA (R+), but not in MSA (R−) or control samples. **B**, Immunohistochemical analysis of BIN1 for MSA (R+). BIN1-positive cells were observed in cerebellar white matter lesions. High-magnification imaging showed that glial cytoplasmic inclusions (GCIs) were positive for BIN1, whereas neurons were negative. **C**, Immunohistochemical analysis of BIN1 in MSA (R− BIN1-immunopositivity was observed uniformly in cerebellar white matter lesions. High-magnification images showed that both GCIs and neurons were negative for BIN1. **D, E**, Fluorescent immunohistochemistry of BIN1, oligodendroglial marker Olig2, and phosphorylated α-synuclein (pSyn). BIN1 colocalized with Olig2 and pSyn. **F**, BIN1-positive GCIs were more frequently observed in the cerebellum of MSA (R+) patients than in those of MSA (R−). Scale Bars: Red = 100 μm, Black = 20 μm, White = 20 μm.

Next, we performed an immunohistochemical analysis to evaluate the localization of BIN1 and identified BIN1-positive cells in the cerebellar white matter lesions of patients with MSA (R+) (Fig. 3B). High-magnification images revealed that the BIN1-positive cells were GCIs, but not neurons (Fig. 3B). Nevertheless, among patients with MSA (R−), BIN1-immunoreactivity was uniformly observed in cerebellar white matter lesions. Both GCIs and neurons were negative for BIN1 (Fig. 3C). Ten patients with Lewy pathology did not demonstrate that Lewy bodies had any BIN-1-immunoreactivities (Supplementary Fig. S4 and Supplementary Table S3). Immunofluorescence showed that BIN1-I colocalized with GCIs stained by the oligodendroglia marker Olig2 (Fig. 3D) and phosphorylated α-synuclein (pSyn) (Fig. 3E and Supplementary Fig. S5). The frequency of BIN1-positive GCIs was significantly higher in the cerebellum of patients with MSA (R+) (23.3 ± 3.2 /mm^2^) than those with patients with MSA (R−) (4.7 ± 2.0 /mm^2^) (*p* = 0.0159) (Fig. 3F). We also performed immunostaining for pSyn but found no difference in the density of GCIs between MSA (R+) and MSA (R−) (Supplementary Fig. S3C, D).

### Fluorescent immunohistochemistry for BIN1 aggregation

In MSA (R+), well-developed pSyn aggregates were observed in the white matter layer, whereas their development was limited in the granule cell layer. In contrast, in MSA (R−), pSyn aggregates were more prominent in the granule cell layer than in the white matter layer. The aggregation of BIN1 was observed in both MSA samples but not in the healthy sample. In the granule cell layer, BIN1 was partially co-localized with pSyn aggregates in both MSA (R+) and MSA (R−) (Fig. 4B). However, the pattern of BIN1 aggregation in the white matter differed between the two groups and was dependent on the repeat expansion. In MSA (R+), larger BIN1-GCIs were observed, co-localized with pSyn aggregates, whereas smaller inclusions were prominent in MSA (R−) (Fig. 4A). Immunofluorescent analysis revealed that BIN1 was embedded in phosphorylated α-synuclein signals in both MSA (R−) and MSA (R+) (Fig. 4C, D), whereas immuno-electron microscopy analysis showed that BIN1 was attached to α-synuclein fibrils via membranous structures in both MSAs (Fig. 4E, F).

**Fig. 4.**
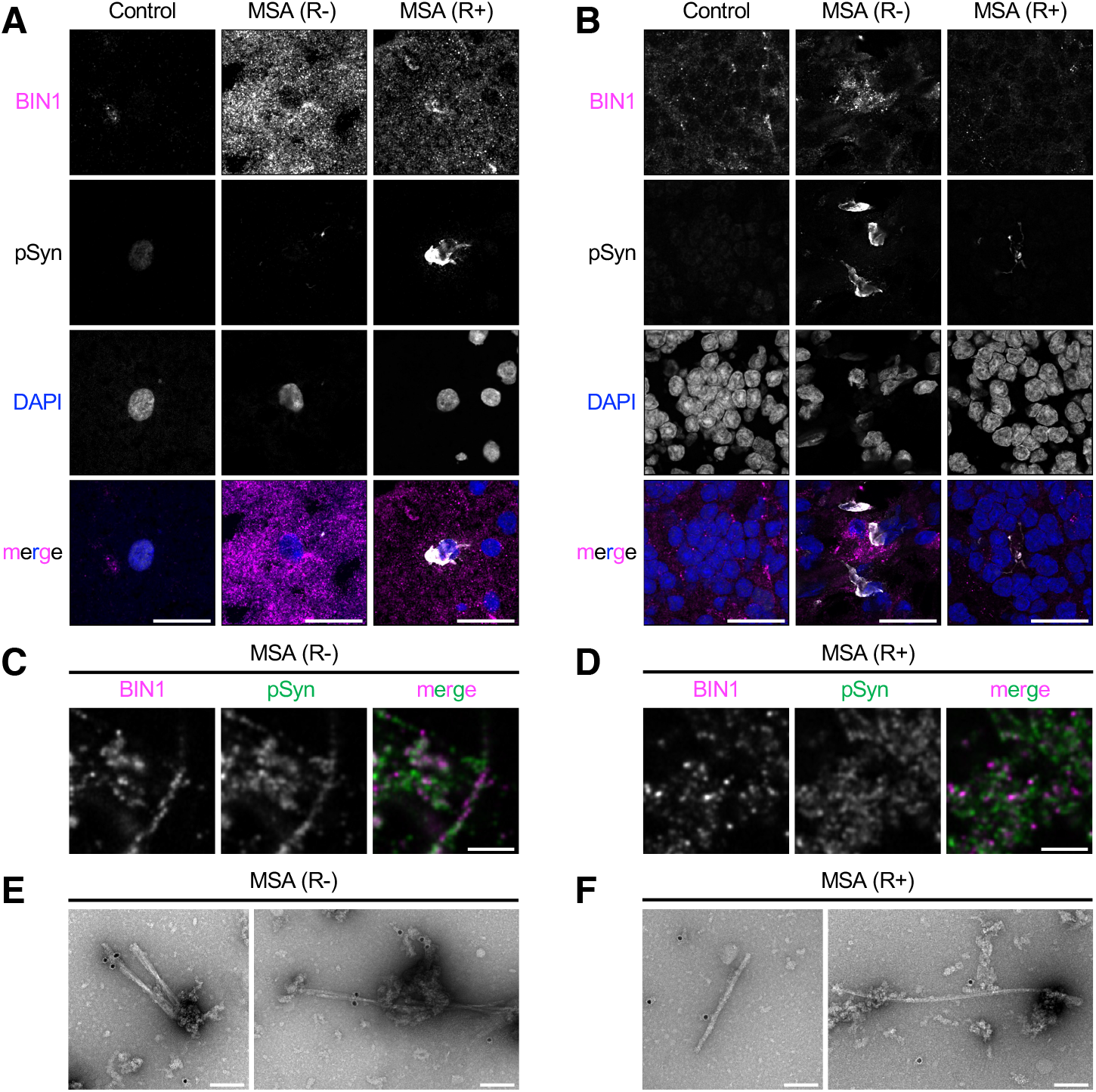
Fluorescent Immunohistochemistry of the cerebellum and immuno-electron microscopy analysis of the sarkosyl-insoluble fraction. **A, B**, Fluorescent immunohistochemistry of BIN1 (magenta), phosphorylated α-synuclein (white), and DAPI (blue) in the white matter (A) and granular cell layers (B) of the cerebellum from control, MSA (R−), and MSA (R+) patients. BIN1-positive GCIs were more frequently observed in the cerebellar white matter lesions of MSA (R+) patients than those of MSA (R−) (A). In the granular cell layer, BIN1 was rarely positive in GCIs in MSA (R−) patients (B). Scale bars: 20 µm. **C, D**, Immunofluorescent analysis of BIN1 (magenta), and phosphorylated α-synuclein (green) in the sarkosyl-insoluble fraction from MSA (R−) (C) and MSA (R+) (D). Scale bars: 1 µm. **E, F**, Immunonegative stain micrographs of filaments in the sarkosyl-insoluble fraction from MSA (R−) (E) and MSA (R+) (F) using an antibody specific for BIN1. Immunogold particles for BIN1 were closely associated with filaments. Scale bars: 100 nm.

### Immunoblot analysis of the brain in MSA with and without the repeat expansion

Next, we conducted immunoblot analysis using brain tissues from control subjects (N = 9) and individuals with MSA (R−) (N = 10) and MSA (R+) (N = 8). Isoform1 of BIN1 (the neuron-specific isoform) in the sarkosyl-soluble fraction was reduced in both MSA (R−) (*p* = 0.05) and MSA (R+) groups (*p* = 0.04) compared to the control group, although the total BIN1 levels remained unchanged (Fig. 5A, C and Supplementary Fig. S6). In the sarkosyl-insoluble fraction, total BIN1 was significantly increased in the MSA (R−) group compared to the control group (*p* < 0.001). However, no significant change was observed in the insoluble total BIN1 levels in the MSA (R+) group compared to those in the control group (*p* = 0.86), despite some individuals displaying elevated levels (Fig. 5B, D and Supplementary Fig. S6). The reduced isoform1 of BIN1 may reflect a reduction in neuronal cells, whereas the increased total BIN1 in the insoluble fraction likely corresponds to the aggregation of BIN1 in oligodendrocytes, as observed in histopathological analysis.

**Fig. 5.**
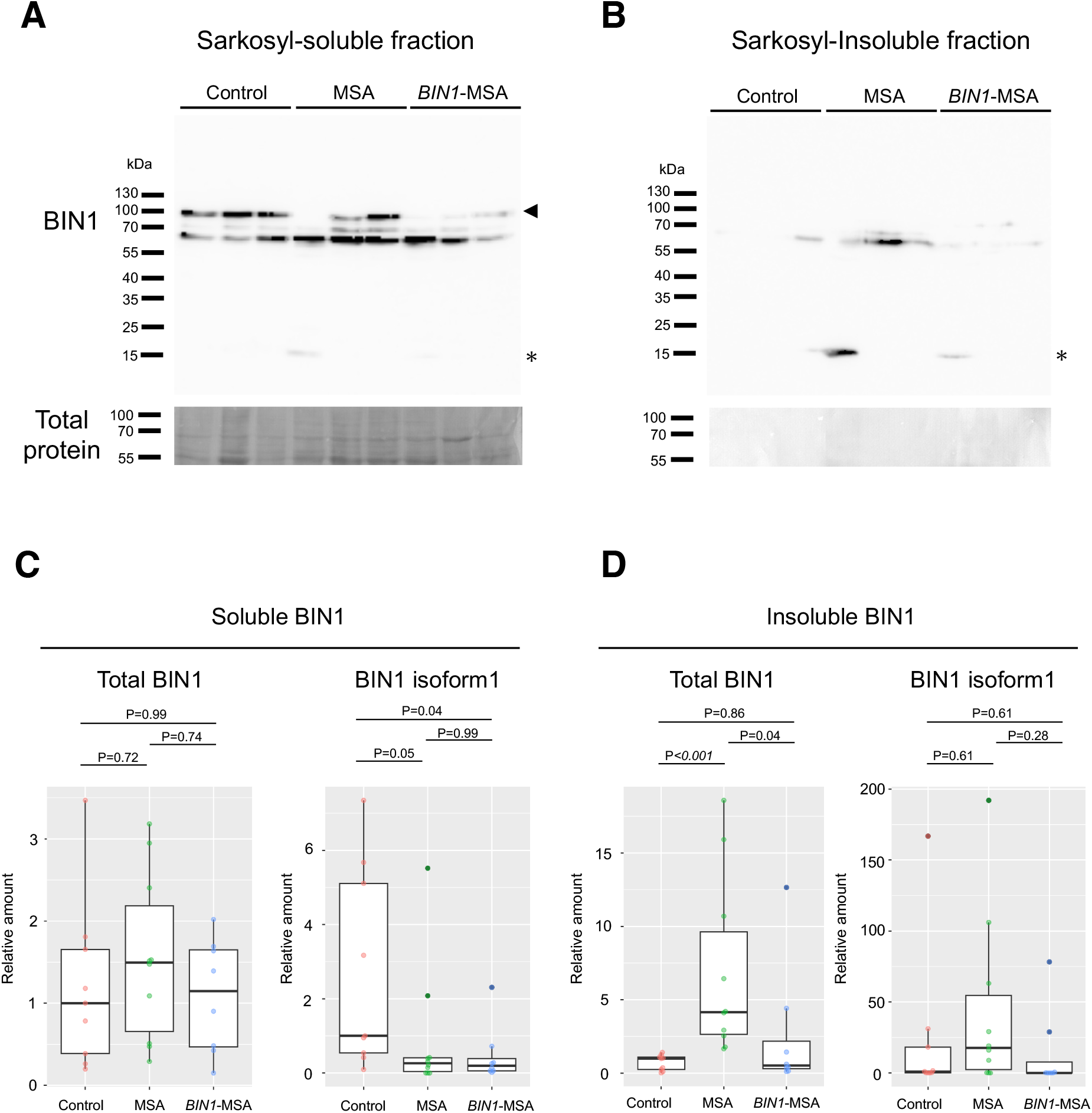
Immunoblot analysis of BIN1 in MSA brain. **A, B**, Representative immunoblots showing BIN1 in the frontal cortex from control subjects (N = 3), MSA (R−) (N = 3), and MSA (R+) (N = 3) in the sarkosyl-soluble fraction (A) and sarkosyl-insoluble fraction (B). The arrowhead indicates the bands corresponding to isoform1 of BIN1. Non-specific bands are marked with an asterisk. **C, D**, show a quantitative analysis of BIN1 protein levels in sarkosyl-soluble (C) and insoluble (D) fractions from control subjects (N = 9), MSA (R−) (N = 10), and MSA (R+) (N = 8). Soluble BIN1 levels are normalized to total protein levels, and insoluble BIN1 levels are quantified per 100 mg of brain tissue. Data are expressed as relative protein quantity compared to the median of the control group. Outliers are highlighted in a dark color. *p*-values were calculated using the Steel–Dwass test.

## Discussion

This study demonstrated three key findings in MSA: (i) expansion of BIN1 repeats, (ii) aggregation of BIN1 with localization to GCIs, and (iii) attachment of BIN1 to phosphorylated α-synuclein filaments in association with membrane-like structures. Taken together, these observations indicate that BIN1 is not merely a genetic risk factor but is actively incorporated into oligodendrocyte-specific pathological structures (GCIs), where it physically interacts with α-synuclein aggregates. Moreover, the presence of membrane-like fragments accompanying these interactions strongly suggests that disruption of BIN1’s intrinsic membrane-remodeling function contributes directly to GCI formation.

Two possible molecular mechanisms through which the *BIN1* repeat expansion may contribute to MSA development are proposed. The first hypothesis is that the abnormal repeat RNA may lead to alpha-synuclein aggregation. RNA binds to alpha-synuclein and promotes its aggregation^14,15^, a hallmark of MSA pathology. The second hypothesis posits that the repeat expansion may disrupt the function of BIN1 through aberrant RNA expression or reduction in BIN1 mRNA. Since BIN1 is important for oligodendrocyte function, its dysfunction may lead to the myelin injury identified in patients with MSA (R+), as previously reported^16^.

BIN1 constitutes part of GCI both in MSA (R+) and MSA (R−) cases. BIN1 aggregation was detected, irrespective of repeat expansion status. BIN1 was associated with fibrillar α-synuclein, accompanied by membranous structures. These membranes likely originate from organelles—such as peroxisomes^17^ and other vesicular compartments^18^—that contribute to GCI composition. The ability of BIN1 to bind to membranes and stabilize curved structures via its BAR domain suggests a role in membrane dynamics^13^. As BIN1 is highly expressed in oligodendrocytes^16^, it may contribute to the formation of glial cytoplasmic inclusions (GCIs) through its involvement in membrane remodeling processes. BIN1 aggregation is a potential diagnostic marker and a promising therapeutic target in MSA.

Our findings suggest that somatic repeat expansions in the brain contribute to the pathogenesis of MSA, potentially explaining its sporadic nature. Somatic copy number variants of *SNCA* contribute to the pathogenesis of MS^19,20^, and somatic repeat expansion in *HTT* causes neurodegeneration in Huntington’s disease^21^. The lack of significant differences in BIN1 repeat expansion rates between clinical MSA samples (blood) and controls (blood) may partly stem from the low diagnostic accuracy of clinical MSA diagnosis. However, it may more strongly indicate that repeat expansion at the lesion site is crucial for MSA onset, while expansion at other sites (such as blood) is not directly related, and that plus, the instability of the expanded repeats might be reducing the correlation between repeat length in peripheral blood and repeat length in the brain.

In conclusion, we identified the AGG repeat expansion in the first intron of *BIN1* in patients with MSA and demonstrated that BIN1 aggregation contributes to MSA pathogenesis, regardless of the presence of the repeat expansion. Our study provides novel insights into the genetic and pathophysiological mechanisms underlying MSA, offering potential targets for the development of therapeutics aimed at this devastating neurodegenerative disorder.

## Materials and Methods

### Study participants

We included an MSA family, 224 patients with clinically diagnosed MSA, and 574 control participants in the Hiroshima University cohort. We obtained brain samples from 67 patients with pathological MSA and 65 control subjects in the Aichi Medical University cohort, as well as from 6 patients with pathological MSA in the Kansai Medical University cohort. Neurologists evaluated all the participants with clinical MSA, whereas neuropathologists diagnosed all brain samples.

Ethical approval was obtained from the ethics committee of Hiroshima University, Aichi Medical University, Kansai Medical University, and Hokkaido University. All participants provided written informed consent, and the study procedures adhered to the tenets of the Declaration of Helsinki.

### Whole genome sequencing

Library preparation was performed using 1.5 μg of DNA sample from a proband individual in the MSA family and a ligation sequencing kit (SQK-LSK-110, Oxford Nanopore Technologies) following the manufacturer’s protocol. Sequencing was conducted using a PromethION and R9.4.1 flow cell (Oxford Nanopore Technologies). Data analysis was performed as previously reported^22^. Briefly, LAST^23^ (version 983, https://github.com/mcfrith/last-genome-alignments) was used for mapping, and tandem genotypes^10^ (https://github.com/mcfrith/tandem-genotypes) were employed to evaluate repeat lengths.

### Genetic screening

Repeat-primed PCR and flanking PCR were performed using TaKaRa LA Taq with the GC Buffer kit (TAKARA) and 7-deaza-dGTP (New England Biolabs Inc.). The detailed protocol and primer sequences are included in Supplementary Table S4. The PCR products were electrophoresed using a 3130xl DNA Genetic analyzer (Applied Biosystems), and fragment analysis was performed using GeneMapper Software (version 5, Applied Biosystems).

### Cas9-mediated enrichment sequencing

Cas9-mediated enrichment sequencing was conducted according to the protocol provided by Oxford Nanopore Technologies Ltd. We used two crRNAs with the target sequences are 5′-GTGCACCCCACGCATTCCGG-3′ and 5′-TGCAACCACGTGATTGCTGG-3′, respectively.

### Histochemical and immunohistochemical analysis

We conducted a histopathological analysis of the cerebellum from four patients with MSA and an AGG repeat expansion in *BIN1*, five with MSA without repeat expansions in *BIN1*, and two control individuals without ataxia or cerebellar atrophy. Postmortem diagnoses were made through routine histochemistry and immunohistochemical analysis at the. Supplementary Table S2 summarizes of the cohort of patients with MSA. We conducted histopathological analysis of the cerebellum, pons, and frontal lobe of one patient with MSA with the repeat expansion. The postmortem diagnosis for this patient was performed using routine histochemistry and immunohistochemical analysis. For each case, 4 μm transverse sections were subjected to immunohistochemical and immunofluorescence analyses. First, the cerebellum was evaluated using histochemical staining methods, including hematoxylin–eosin, Klüver–Barrera (KB), and Gallyas–Braak silver (GB) stains.

For immunohistochemical analysis, endogenous peroxidase activity was blocked by incubating the sections in 0.3% H2O2 in absolute methanol for 20 min. After washing in phosphate-buffered saline (PBS), the sections were incubated overnight at 4 °C with primary antibodies. Supplementary Table S5 lists the primary antibodies used. Antibody binding was visualized using horseradish peroxidase (HRP)-labeled goat anti-mouse or anti-rabbit antibodies (1:100, DAKO, Glostrup, Denmark) for 30 min at 20 °C. After washing the sections three times in PBS, they were incubated with 3,3′-diaminobenzidine (DAB; DAKO) to visualize the localization of epitopes. Nuclear counterstaining was performed using DAPI. The sections were photographed using a BIOREVO BZ-700 fluorescence microscope (Keyence) and a BX43 Olympus microscope with a DP28 digital camera and CellSens standard software (Olympus). For quantitative analysis, we examined 20 serial fields of cerebellar white matter lesions at 200× magnification to calculate the densities of glial cytoplasmic inclusions (GCIs).

Lewy pathology was evaluated based on our previous report^24^. The HE sections of the pons were photographed using a BX43 Olympus microscope with a DP28 digital camera and CellSens standard software (Olympus). The sections were decolorized in 70% ethanol for 24 h and immunostained.

For immunofluorescence analysis, sections were incubated with primary mouse and rabbit antibodies overnight at 4°C, followed by washing in PBS. Supplementary Table S5 lists the primary antibodies used. The sections were then visualized using anti-mouse secondary antibody conjugated with Alexa Fluor 568 and anti-rabbit secondary antibody conjugated with Alexa Fluor 488. The primary antibodies consisted of mouse monoclonal antibodies against Olig2 and pSyn and rabbit polyclonal antibodies against BIN1-I (Supplementary Table S5). Nuclear counterstaining was performed using DAPI (Nacalai Tesque). The sections were photographed using a BIOREVO BZ-700 fluorescence microscope (Keyence).

For the detection of BIN1 aggregations using immunofluorescent analysis, cryosections of frozen cerebellar samples mounted on glass slides were air-dried and fixed in 95% ethanol at 4 °C for 30 min, followed by 100% acetone at room temperature (RT) for 1 min. The sections were rinsed in PBS (−) three times for 5 min, blocked with 1% BSA/PBS (−) at RT for 3 h, and incubated with primary antibodies against pSyn and BIN1-I (Supplementary Table S5) at RT for 3 h. After washing with PBS (−) three times, the sections were incubated with DAPI and secondary antibodies (anti-mouse secondary antibody conjugated with Alexa Fluor 594 and anti-rabbit secondary antibody conjugated with Alexa Fluor 647) at RT for 2 h. The sections were washed three times with PBS (−) and mounted with Prolong Diamond antifade medium (Thermo Fisher Scientific). The stained samples were examined using an LSM800 confocal microscope (Carl Zeiss).

### Immunoblot analysis for brain samples

Protein extraction from brain tissues was performed as previously reported^25^. Briefly, 200 mg of brain tissue was homogenized in 4 mL of extraction buffer (10 mM Tris-HCl, pH 7.5, containing 10% sucrose, 0.8 M NaCl, 1 mM EGTA). Sarkosyl (final concentration: 2%) was added to homogenates, and the mixture was incubated for 30 min. at 37 °C. After centrifugation for 10 min at 10,000 × *g*, the supernatants were further centrifuged at 166,000 × *g* for 20 min at 25 °C. The resulting supernatants were stored as sarkosyl-soluble fractions, with protein concentration quantified using the Pierce BCA Protein Assay Kit (Thermo Fisher). The pellets were washed with saline and ultracentrifuged as described above, and then resuspended in 8 M urea and stored as sarkosyl-insoluble fractions. Protein samples were stored at −80 °C until use. For immunoblotting, 20 μg of sarkosyl-soluble fraction and proteins from the insoluble fraction of 100 mg of brain tissues were separated by sodium dodecyl sulfate-polyacrylamide gel electrophoresis (SDS-PAGE) on a 10% gel. Proteins were transferred to polyvinylidene fluoride (PVDF) membranes (FluoroTrans, 0.2 μm pore size, Pall) using the TransBlot Turbo Transfer System (Bio-Rad). Total protein levels were assessed via Ponceau S staining (Sigma Aldrich).

The membranes were incubated overnight with a primary rabbit polyclonal antibody for BIN1 in 5% skim milk, followed by a 1 h incubation with an anti-rabbit secondary antibody (Supplementary Table S5). Chemiluminescence was detected using Immobilon Forte Western HRP substrate (Merck Millipore). Protein quantification was performed using ImageJ/Fiji software. BIN1 protein levels in the soluble fraction were normalized to total protein levels.

### Immuno-electron Microscopy Analysis

The sarkosyl-insoluble fraction was prepared as described^26^. For immunonegative staining, aliquots of the sarkosyl-insoluble fraction were placed on formvar-coated 200-mesh copper grids (Nisshin EM), fixed in 4% PFA/PBS (Nacalai Tesque), blocked with 2% BSA/PBS (−), and incubated with the primary antibody against BIN1-N (Supplementary Table S5) at RT for 2 h. After washing with PBS (−), the grids were incubated with secondary antibody conjugated to 12 nm gold particles (Supplementary Table S5) at RT for 1 h. The grids were then stained with 1% uranyl acetate. Images were acquired using a 120 kV FEI Tecnai G2 Spirit Bio TWIN electron microscope equipped with an Emsis PHURONA CMOS camera.

### Statistics

All statistical analyses were performed using R software (version 4.0.3) or Prism 8 software (GraphPad Software). A *p*-value of <0.05 was considered statistically significant. Fisher’s exact tests were used to assess genetic association using R software, Mann–Whitney *U* tests were performed for immunohistochemical analysis using Prism 8 software, and Steel– Dwass tests were used for immunoblot analysis with R software.

## Data Availability

The data that support the findings of this study are available from the corresponding author upon reasonable request.

## Acknowledgments

We acknowledge Professor Hidefumi Ito for critical reading of the manuscript and constructive comments. We also thank Ms. Eiko Nakajima, Ms. Rika Tai (Hiroshima University), Mr. Hiroki Fujisawa, Ms. Arisa Kan (National Hospital Organization Kure Medical Center and Chugoku Cancer Center, Kure, Japan), and Dr. Azumi Yoshimura (Institute of Biomedical Research and Education, Science Research Center, Yamaguchi University) for their technical assistance.

## Author contributions

Conceptualization: HK, KK, and TK

Methodology: TA, AA, Yasuhi I, MN, Mai K, Masaki K, Yuishin I, and IY

Investigation: KK, TK, and TI

Supervision: HK, KK, and TK

Writing – original draft: HK, KK, TK, TI, and TM

Writing – review & editing: TI, TM, AT, and KM

## Supplementary Materials

Figs. S1 to S6

Tables S1 to S5

## References and Notes

1. N. Stefanova, G. K. Wenning, Multiple system atrophy: at the crossroads of cellular, molecular and genetic mechanisms. Nat. Rev. Neurosci. 24, 334–346 (2023).

2. M. I. Papp, J. E. Kahn, P. L. Lantos, Glial cytoplasmic inclusions in the CNS of patients with multiple system atrophy (striatonigral degeneration, olivopontocerebellar atrophy and Shy-Drager syndrome). J. Neurol. Sci. 94, 79–100 (1989).

3. Y. Nakazato, H. Yamazaki, J. Hirato, Y. Ishida, H. Yamaguchi, Oligodendroglial microtubular tangles in olivopontocerebellar atrophy. J. Neuropathol. Exp. Neurol. 49, 521–530 (1990).

4. K. Wakabayashi, M. Yoshimoto, S. Tsuji, H. Takahashi, Alpha-synuclein immunoreactivity in glial cytoplasmic inclusions in multiple system atrophy. Neurosci. Lett. 249, 180–182 (1998).

5. S.W. Scholz, H. Houlden, C. Schulte, M. Sharma, A. Li, D. Berg, A. Melchers, R. Paudel, J. R. Gibbs, J. Simon-Sanchez, C. Paisan-Ruiz, J. Bras, J. Ding, H. Chen, B. J. Traynor, S. Arepalli, R. R. Zonozi, T. Revesz, J. Holton, N. Wood, A. Lees, W. Oertel, U. Wüllner, S. Goldwurm, M. T. Pellecchia, T. Illig, O. Riess, H. H. Fernandez, R. L. Rodriguez, M. S. Okun, W. Poewe, G. K. Wenning, J. A. Hardy, A. B. Singleton, F. Del Sorbo, S. Schneider, K. P. Bhatia, T. Gasser, SNCA variants are associated with increased risk for multiple system atrophy. Ann. Neurol. 65, 610–614 (2009).

6. M. G. Heckman, L. Schottlaender, A. I. Soto-Ortolaza, N. N. Diehl, S. Rayaprolu, K. Ogaki, S. Fujioka, M. E. Murray, W. P. Cheshire, R. J. Uitti, Z. K. Wszolek, M. J. Farrer, A. Sailer, A. B. Singleton, P. F. Chinnery, M. J. Keogh, S. M. Gentleman, Janice L Holton 1, Kiely Aoife, D.M.A. Mann, S. Al-Sarraj, C. Troakes, D. W. Dickson, H. Houlden, O. A. Ross, LRRK2 exonic variants and risk of multiple system atrophy. Neurology 83, 2256–2261 (2014).

7. Multiple-System Atrophy Research Collaboration, Mutations in COQ2 in familial and sporadic multiple-system atrophy. N. Engl. J. Med. 369, 233–244 (2013).

8. F. Hopfner, A. K. Tietz, V. C. Ruf, O. A. Ross S. Koga, D. Dickson, A. Aguzzi, J. Attems, T. Beach, A. Beller, W. P. Cheshire, V. van Deerlin, P. Desplats, G. Deuschl, C. Duyckaerts, D. Ellinghaus, V. Evsyukov, M. E. Flanagan, A. Franke, M. P. Frosch, M. Gearing, E. Gelpi, J. A. van Gerpen, B. Ghetti, J.D. Glass, L. T. Grinberg, G. Halliday, I. Helbig, M. Höllerhage, I. Huitinga, D.J. Irwin, D. C. Keene, G. G. Kovacs, E. B. Lee, J. Levin, M.J. Martí, I. Mackenzie, I. McKeith, C. Mclean, B. Mollenhauer, M. Neumann, K. L. Newell, A. Pantelyat, M. Pendziwiat, A. Peters, L. M. Porcel, A. Rabano, R. Matěj, A. Rajput, A. Rajput, R. Reimann, W. K. Scott, W. Seeley, S. Selvackadunco, T. Simuni, C. Stadelmann, P. Svenningsson, A. Thomas, C. Trenkwalder, C. Troakes, J. Q. Trojanowski, R. J. Uitti, C. L. White, Z. K. Wszolek, T. Xie, T. Ximelis, J. Yebenes; Alzheimer’s Disease Genetics Consortium; U. Müller, G. D. Schellenberg, J. Herms, G. Kuhlenbäumer, G. Höglinger, Common variants near ZIC1 and ZIC4 in autopsy-confirmed multiple system atrophy. Mov. Disord. 37, 2110–2121 (2022).

9. R. Chia, R. Chia, A. Ray, Z. Shah, J. Ding, P. Ruffo, M. Fujita, V. Menon, S. Saez-Atienzar, P. Reho, K. Kaivola, R. L. Walton, R. H. Reynolds, R. Karra, S. Sait, F. Akcimen, M. Diez-Fairen, I. Alvarez., A. Fanciulli, N. Stefanova, K. Seppi, S. Duerr, F. Leys, F. Krismer., V. Sidoroff, A. Zimprich, W. Pirker, O. Rascol, A. Foubert-Samier, W. G. Meissner, F. Tison, A. Pavy-Le Traon, M. T. Pellecchia, P. Barone, M. C. Russillo, J. Marín-Lahoz, J. Kulisevsky, S. Torres, P. Mir 21, Maria T. Periñán, C. Proukakis, V. Chelban, L. Wu, Y. Y. Goh, L. Parkkinen, M. T. Hu, C. Kobylecki, J. A. Saxon, S. Rollinson, E. Garland, I. Biaggioni, I. Litvan, I. Rubio, R. N. Alcalay, K. T. Kwei, S. J. Lubbe, Q. Mao, M. E. Flanagan, R. J. Castellani, V. Khurana, A. Ndayisaba, A. Calvo, G. Mora, A. Canosa, G. Floris, R. C. Bohannan, A. Moore, L. Norcliffe-Kaufmann, J.-A. Palma, H. Kaufmann, C. Kim, M. Iba, E. Masliah, T. Dawson, L. S. Rosenthal, A. Pantelyat, M. S. Albert, O. Pletnikova, J. C. Troncoso, J. Infante, C. Lage, P. Sánchez-Juan, G. E. Serrano, T. G. Beach, P. Pastor, H. R. Morris, D. Albani, J. Clarimon, G. K. Wenning, J. A. Hardy, M. Ryten, E. Topol, A. Torkamani, A. Chiò, D. A. Bennett, P. L. De Jager, P. A. Low, W. Singer, W. P. Cheshire, Z. K. Wszolek, D. W. Dickson, B. J. Traynor, J. R. Gibbs, C. L. Dalgard, O. A. Ross, H. Houlden, S. W. Scholz, Genome sequence analyses identify novel risk loci for multiple system atrophy. Neuron 112, 2142–2156.e5 (2024).

10. S. Mitsuhashi, M. C. Frith, T. Mizuguchi, S. Miyatake, T. Toyota, H. Adachi, Y. Oma, Y. Kino, H. Mitsuhashi, N. Matsumoto, Tandem-genotypes: robust detection of tandem repeat expansions from long DNA reads. Genome Biol. 20, 1–17 (2019).

11. A.-S. Nicot, A. Toussaint, V. Tosch, C. Kretz, C. Wallgren-Pettersson, E. Iwarsson, H. Kingston, J.-M. Garnier, V. Biancalana, A. Oldfors, J.-L. Mandel, J. Laporte, Mutations in amphiphysin 2 (BIN1) disrupt interaction with dynamin 2 and cause autosomal recessive centronuclear myopathy. Nat. Genet. 39, 1134– 1139 (2007).

12. C. Bellenguez, F. Küçükali, I. E. Jansen, L. Kleineidam, S. Moreno-Grau, N. Amin, A. C. Naj, R. Campos-Martin, B. Grenier-Boley, V. Andrade, P. A. Holmans, A. Boland, V. Damotte, S. J. van der Lee, M. R. Costa, T. Kuulasmaa, Q. Yang, I. de Rojas, J. C. Bis, A. Yaqub, I. Prokic, J. Chapuis, S. Ahmad, V. Giedraitis, D. Aarsland, P. Garcia-Gonzalez, C. Abdelnour, E. Alarcón-Martín, D. Alcolea, M. Alegret, I. Alvarez, V. Álvarez, N. J. Armstrong, A. Tsolaki, C. Antúnez, I. Appollonio, M. Arcaro, S. Archetti, A. A. Pastor, B. Arosio, L. Athanasiu, H. Bailly, N. Banaj, M. Baquero, S. Barral, A. Beiser, A. Belén Pastor, J.E. Below, P. Benchek, L. Benussi, C. Berr, C. Besse, V. Bessi, G. Binetti, A. Bizarro, R. Blesa, M. Boada, E. Boerwinkle, B. Borroni, S. Boschi, P. Bossù, G. Bråthen, J. Bressler, C. Bresner, H. Brodaty, K. J. Brookes, L. I. Brusco, D. Buiza-Rueda, K. Bûrger, V. Burholt, W. S. Bush, M. Calero, L. B. Cantwell, G. Chene, J. Chung, M. L. Cuccaro, Á. Carracedo, R. Cecchetti, L. Cervera-Carles, C. Charbonnier, H.-H. Chen, C. Chillotti, S. Ciccone, J. A. H. R. Claassen, C. Clark, E. Conti, A. Corma-Gómez, E. Costantini, C. Custodero, D. Daian, M. C. Dalmasso, A. Daniele, E. Dardiotis, J.-F. Dartigues, P. P. de Deyn, K. de Paiva Lopes, L. D. de Witte, S. Debette, J. Deckert, T. Del Ser, N. Denning, A. DeStefano, M. Dichgans, J. Diehl-Schmid, M. Diez-Fairen, P. D. Rossi, S. Djurovic, E. Duron, E. Düzel, C. Dufouil, G. Eiriksdottir, S. Engelborghs, V. Escott-Price, A. Espinosa, M. Ewers, K.M. Faber, T. Fabrizio, S. Fallgaard Nielsen, David W Fardo, L. Farotti, C. Fenoglio, M. Fernández-Fuertes, R. Ferrari, C. B. Ferreira, E. Ferri, B. Fin, P. Fischer, T. Fladby, K. Fließbach, B. Fongang, M. Fornage, J. Fortea, T. M. Foroud, S. Fostinelli, N. C. Fox, E. Franco-Macías, M. J. Bullido, A. Frank-García, L. Froelich, B. Fulton-Howard, D. Galimberti, J.M. García-Alberca, P. García-González, S. Garcia-Madrona, G. Garcia-Ribas, R. Ghidoni, I. Giegling, G. Giorgio, A. M. Goate, O. Goldhardt, D. Gomez-Fonseca, A. González-Pérez, C. Graff, G. Grande, E. Green, T. Grimmer, E. Grünblatt, M. Grunin, V. Gudnason, T. Guetta-Baranes, A. Haapasalo, G. Hadjigeorgiou, J. L. Haines, K. L. Hamilton-Nelson, H. Hampel, O. Hanon, J. Hardy, A. M Hartmann, L. Hausner, J. Harwood, S. Heilmann-Heimbach, S. Helisalmi, M. T. Heneka, I. Hernández, M. J. Herrmann, P. Hoffmann, C. Holmes, H. Holstege, R. H. Vilas, M. Hulsman, J. Humphrey, G. J. Biessels, X. Jian, C. Johansson, G. R. Jun, Y. Kastumata, J. Kauwe, P. G. Kehoe, L. Kilander, A. Kinhult Ståhlbom, M. Kivipelto, A. Koivisto, J. Kornhuber, M. H. Kosmidis, W. A. Kukull, P. P. Kuksa, B. W. Kunkle, A. B. Kuzma, C. Lage, Erika J Laukka, L. Launer, A. Lauria, C.-Y. Lee, J. Lehtisalo, O. Lerch, A. Lleó, W. Longstreth Jr, O. Lopez, A. L. de Munain, S. Love, M. Löwemark, L. Luckcuck, K. L. Lunetta, Y. Ma, J. Macías, C. A. MacLeod, W. Maier, F. Mangialasche, M. Spallazzi, M. Marquié, R. Marshall, E. R. Martin, A. M. Montes, C. M. Rodríguez, C. Masullo, R. Mayeux, S. Mead, P. Mecocci, M. Medina, A. Meggy, S. Mehrabian, S. Mendoza, M. Menéndez-González, P. Mir, S. Moebus, M. Mol, L. Molina-Porcel, L. Montrreal, L. Morelli, F. Moreno, K. Morgan, T. Mosley, M. M. Nöthen, C. Muchnik, S. Mukherjee, B. Nacmias, T. Ngandu, G. Nicolas, B. G. Nordestgaard, R. Olaso, A. Orellana, M. Orsini, G. Ortega, A. Padovani, C. Paolo, G. Papenberg, L. Parnetti, F. Pasquier, P. Pastor, G. Peloso, A. Pérez-Cordón, J. Pérez-Tur, P. Pericard, O. Peters, Y. A. L. Pijnenburg, J. A. Pineda, G. Piñol-Ripoll, C. Pisanu, T. Polak, J. Popp, D. Posthuma, J. Priller, R. Puerta, O. Quenez, I. Quintela, J. Q. Thomassen, A. Rábano, I. Rainero, F. Rajabli, I. Ramakers, L. M. Real, M. J. T. Reinders, C. Reitz, D. Reyes-Dumeyer, P. Ridge, S. Riedel-Heller, P. Riederer, N. Roberto, E. Rodriguez-Rodriguez, A. Rongve, I. R. Allende, M. Rosende-Roca, J. L. Royo, E. Rubino, D. Rujescu, M. E. Sáez, P. Sakka, I. Saltvedt, Á. Sanabria, M. B. Sánchez-Arjona, F. Sanchez-Garcia, P. S. Juan, R. Sánchez-Valle, S. B. Sando, C. Sarnowski, C. L. Satizabal, M. Scamosci, N. Scarmeas, E. Scarpini, P. Scheltens, N. Scherbaum, M. Scherer, M. Schmid, A. Schneider, J. M. Schott, G. Selbæk, D. Seripa, M. Serrano, J. Sha, A. A. Shadrin, O. Skrobot, S. Slifer, G. J. L. Snijders, H. Soininen, V. Solfrizzi, A. Solomon, Y. Song, S. Sorbi, O. Sotolongo-Grau, G. Spalletta, A. Spottke, A. Squassina, E. Stordal, J. P. Tartan, L. Tárraga, N. Tesí, A. Thalamuthu, T. Thomas, G. Tosto, L. Traykov, L. Tremolizzo, A. Tybjærg-Hansen, A. Uitterlinden, A. Ullgren, I. Ulstein, S. Valero, O. Valladares, C. Van Broeckhoven, J. Vance, B. N. Vardarajan, A. van der Lugt, J. Van Dongen, J. van Rooij, J. van Swieten, R. Vandenberghe, F. Verhey, J.-S. Vidal, J. Vogelgsang, M. Vyhnalek, M. Wagner, D. Wallon, L.-S. Wang, R. Wang, L. Weinhold, J. Wiltfang, G. Windle, B. Woods, M. Yannakoulia, H. Zare, Y. Zhao, X. Zhang, C. Zhu, M. Zulaica; EADB; GR@ACE; DEGESCO; EADI; GERAD; Demgene; FinnGen; ADGC; CHARGE; L.A. Farrer, B. M. Psaty, M. Ghanbari, Raj, P. Sachdev, K. Mather, F. Jessen, M. A. Ikram, A. de Mendonça, J. Hort, M. Tsolaki, M. A. Pericak-Vance, P. Amouyel, J. Williams, R. Frikke-Schmidt, J. Clarimon, J.-F. Deleuze, G. Rossi, S. Seshadri, O. A. Andreassen, M. Ingelsson, M. Hiltunen, K. Sleegers, G. D. Schellenberg, C. M. van Duijn, R. Sims, W. M. van der Flier, A. Ruiz, A. Ramirez, J.-C. Lambert, New insights into the genetic etiology of Alzheimer’s disease and related dementias. Nat. Genet. 54, 412–436 (2022).

13. P. Dourlen, D. Kilinc, I. Landrieu, J. Chapuis, J.-C. Lambert, BIN1 and Alzheimer’s disease: the tau connection. Trends Neurosci. 48, 349–361 (2025).

14. J. Rupert, M. Monti, E. Zacco, G.G. Tartaglia, RNA sequestration driven by amyloid formation: the alpha synuclein case. Nucleic Acids Res. 51, 11466–11478 (2023).

15. K. Matsuo, S. Asamitsu, K. Maeda, H. Suzuki, K. Kawakubo, G. Komiya, K. Kudo, Y. Sakai, K. Hori1, S. Ikenoshita, S. Usuki, S. Funahashi, H. Oizumi, A. Takeda, Y. Kawata, T. Mizobata, N. Shioda, Y. Yabuki, RNA G-quadruplexes form scaffolds that promote neuropathological α-synuclein aggregation. Cell S0092-8674(24)01134–6 (2024) doi:10.1016/j.cell.2024.09.037.

16. D. Rossi, P.V. Buggia-Prévot, B. L. L. Clayton, J. B. Vasquez, C. van Sanford, R. J. Andrew, R. Lesnick, A. Botté, C. Deyts, S. Salem, E. Rao, R. C. Rice, A. Parent, S. Kar, B. Popko, P. Pytel, S. Estus, G. Thinakaran, Predominant expression of Alzheimer’s disease-associated BIN1 in mature oligodendrocytes and localization to white matter tracts. Mol. Neurodegener. 11, 59 (2016).

17. C. Böing, M. Di Fabrizio, D. Burger, J. G. J. M. Bol, E. Huisman, A. J. M. Rozemuller, W. D. J. van de Berg, H. Stahlberg, A. J. Lewis, Distinct ultrastructural phenotypes of glial and neuronal alpha-synuclein inclusions in multiple system atrophy. Brain 147, 3727–3741 (2024).

18. F. Mori, Y. Miki, K. Tanji, T. Kon, M. Tomiyama, A. Kakita, K. Wakabayashi, Role of VAPB and vesicular profiles in α-synuclein aggregates in multiple system atrophy. Brain Pathol. 31, e13001 (2021).

19. D. Perez-Rodriguez, M. Kalyva, M. Leija-Salazar, T. Lashley, M. Tarabichi, V. Chelban, S. Gentleman, L. Schottlaender, H. Franklin, G. Vasmatzis, H. Houlden, A. H. V. Schapira, T. T. Warner, J. L. Holton, Z. Jaunmuktane, C. Proukakis, Investigation of somatic CNVs in brains of synucleinopathy cases using targeted SNCA analysis and single cell sequencing. Acta Neuropathol. Commun. 7, 219 (2019).

20. M. E. Garcia-Segura, D. Perez-Rodriguez, D. Chambers, Z. Jaunmuktane, C. Proukakis, Somatic SNCA copy number variants in multiple system atrophy are related to pathology and inclusions. Mov. Disord. 38, 338–342 (2023).

21. R. E. Handsaker, S. Kashin, N. M. Reed, S. Tan, W.-S. Lee, T. M. McDonald, K. Morris, N. Kamitaki, C. D. Mullally, N. R. Morakabati, M. Goldman, G. Lind, R. Kohli, E. Lawton, M. Hogan, K. Ichihara, S. Berretta, S. A. McCarroll, Long somatic DNA-repeat expansion drives neurodegeneration in Huntington’s disease. Cell 188, 623–639.e19 (2025).

22. K. Kume, T. Kurashige, K. Muguruma, H. Morino, Y. Tada, M. Kikumoto, T. Miyamoto, S. N. Akutsu, Y. Matsuda, S. Matsuura, M. Nakamori, A. Nishiyama, R. Izumi, T. Niihori, M. Ogasawara, N. Eura, T. Kato, M. Yokomura, Y. Nakayama, H. Ito, M. Nakamura, K. Saito, Y. Riku, Y. Iwasaki, H. Maruyama, Y. Aoki, I. Nishino, Y. Izumi, M. Aoki, H. Kawakami, CGG repeat expansion in LRP12 in amyotrophic lateral sclerosis. Am. J. Hum. Genet. 110, 1086–1097 (2023).

23. S. M. Kielbasa, R. Wan, K. Sato, P. Horton, M. C. Frith, Adaptive seeds tame genomic sequence comparison. Genome Res. 21, 487–493 (2011).

24. T. Kurashige, T. Takahashi, Y. Yamazaki, M. Hiji, Y. Izumi, T. Yamawaki, M. Matsumoto, Localization of CHMP2B-immunoreactivity in the brainstem of Lewy body disease. Neuropathology 33, 237–245 (2013).

25. A. Tarutani, T. Arai, S. Murayama, S. Hisanaga, M. Hasegawa, Potent prion-like behaviors of pathogenic α-synuclein and evaluation of inactivation methods. Acta Neuropathol. Commun. 6, 29 (2018).

26. M. Schweighauser, Y. Shi, A. Tarutani, F. Kametani, A. G. Murzin, B. Ghetti, T. Matsubara, T. Tomita, T. Ando, K. Hasegawa, S. Murayama, M. Yoshida, M. Hasegawa, S. H.W. Scheres, M. Goedert, Structures of α-synuclein filaments from multiple system atrophy. Nature 585, 464–469 (2020).

