## Supplementary material for "AGG Repeat Expansion and Aggregation of BIN1 in Multiple System Atrophy": Figs. S1 to S6. Tables S1 to S5

**The PDF file includes:**

Figs. S1 to S6
Tables S1 to S5

257 repeats

$(AGG)_{68}(AGGG)_{1}(AGG)_{70}(AG)_{1}(AGG)_{102}(AG)_{1}(AGG)_{3}(AGGG)_{1}(AGG)_{7}(AGGGG)_{1}(AGG)_{2}$

AGGAGGAGGAGGAGGAGGAGGAGGAGGAGGAGGAGGAGGAGGAGGAGGAGGAGGAGGAGGAGGAGGAGGAGGAGGAGGAGGAGGAGGAGGAGGAGGAGGAGGAGG
AGGAGGAGGAGGAGGAGGAGGAGGAGGAGGAGGAGGAGGAGGAGGAGGAGGAGGAGGAGGAGGAGGAGGAGGAGGAGGAGGAGGAGGAGGAGGAGGAGGAGGG
AGGAGGAGGAGGAGGAGGAGGAGGAGGAGGAGGAGGAGGAGGAGGAGGAGGAGGAGGAGGAGGAGGAGGAGGAGGAGGAGGAGGAGGAGGAGGAGGAGGAGGAGG
AGGAGGAGGAGGAGGAGGAGGAGGAGGAGGAGGAGGAGGAGGAGGAGGAGGAGGAGGAGGAGGAGGAGGAGGAGGAGGAGGAGGAGGAGGAGGAGGAGGAGGAGG
AGAGGAGGAGGAGGAGGAGGAGGAGGAGGAGGAGGAGGAGGAGGAGGAGGAGGAGGAGGAGGAGGAGGAGGAGGAGGAGGAGGAGGAGGAGGAGGAGGAGGAGG
AGGAGGAGGAGGAGGAGGAGGAGGAGGAGGAGGAGGAGGAGGAGGAGGAGGAGGAGGAGGAGGAGGAGGAGGAGGAGGAGGAGGAGGAGGAGGAGGAGGAGGAGG
AGGAGGAGGAGGAGGAGGAGGAGGAGGAGGAGGAGGAGGAGGAGGAGGAGGAGGAGGAGGAGGAGGAGGAGGAGGAGGAGGAGGAGGAGGAGGAGGAGAGG
AGGAGGAGGGAGGAGGAGGAGGAGGAGGAGGAGGGGAGGAGG

**Fig. S1. Consensus sequence of AGG repeat in the proband individual.** Adenine and guanine are highlighted in yellow and light blue, respectively.

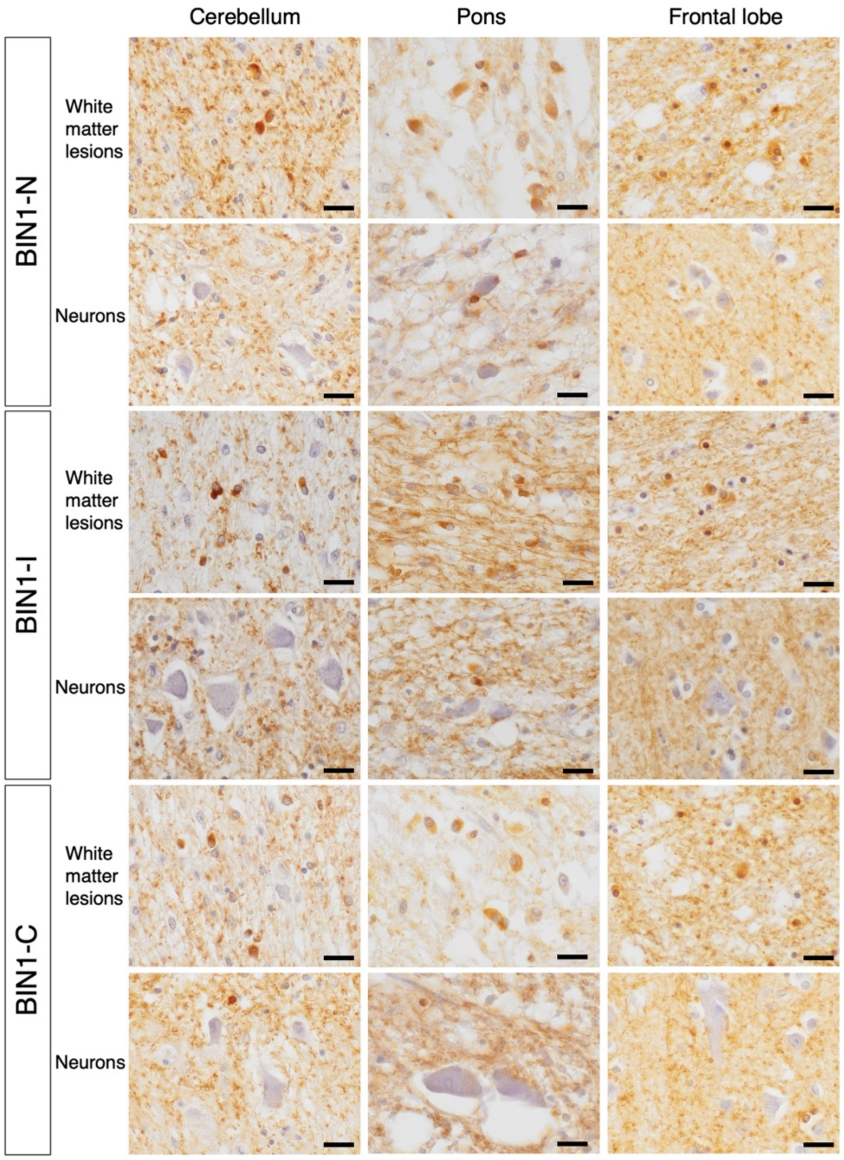


**Fig. S2. Pathological analysis of MSA with somatic repeat expansion in *BIN1*.** BIN1 immunostaining revealed that GCIs exhibit immunoreactivity against BIN1, whereas neurons do not. Scale Bars: 20µm.

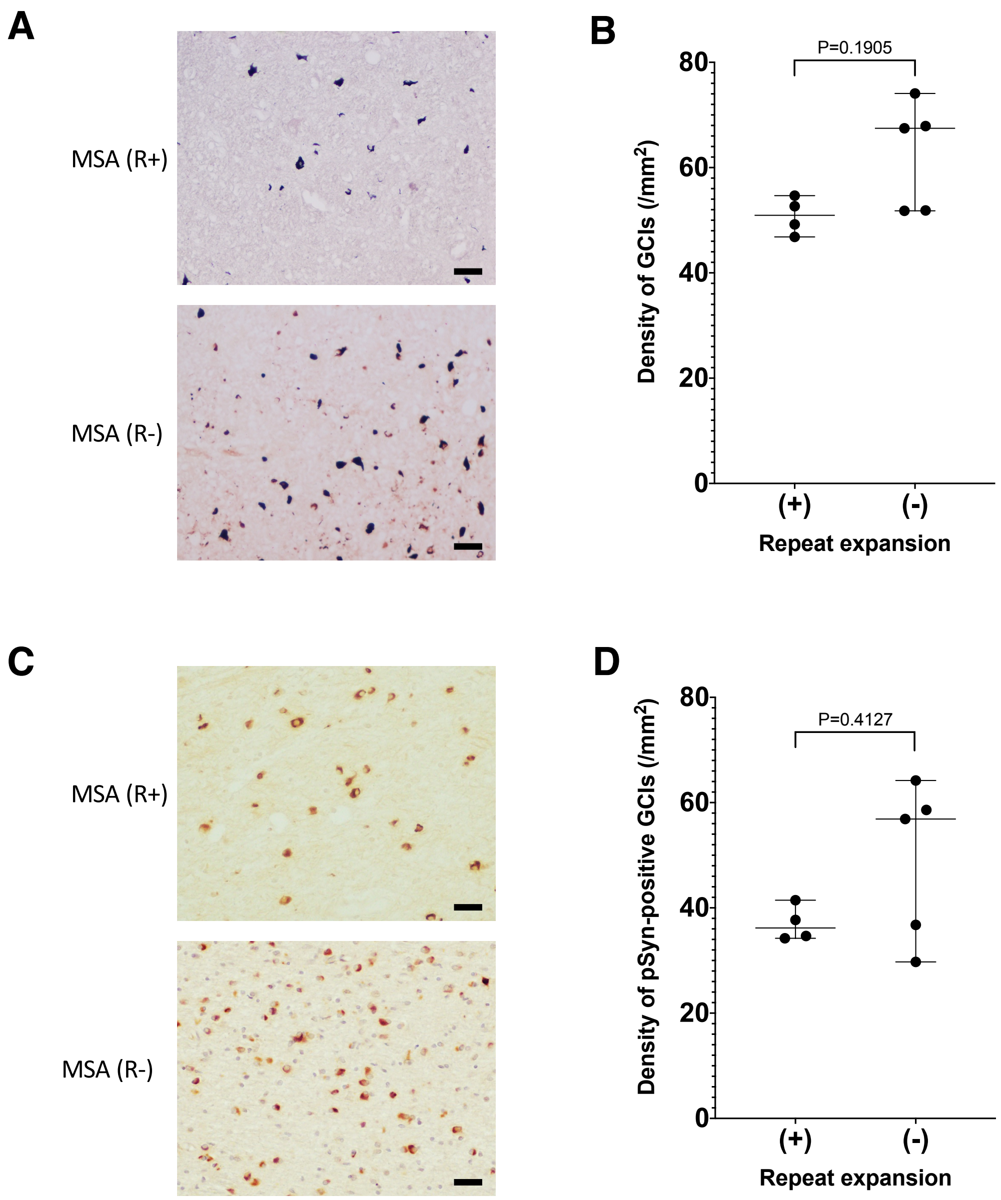


**Fig. S3. Glial cytoplasmic inclusions stained with Gallyas–Braak silver staining and antibody against phosphorylated α-synuclein. A,** Glial cytoplasmic inclusions (GCIs) stained with Gallyas–Braak (GB) silver staining. **B,** Quantification of the density of GCIs stained with GB. No significant differences in the density of GCIs in the cerebellar white matter lesions were observed between MSA with and without repeat expansion. **C,** GCIs stained with phosphorylated α-synuclein (pSyn). **D**, the density of pSyn-positive GCIs. No significant difference in the density of GCIs stained with pSyn was observed in the cerebellar white matter lesions between MSA with and without repeat expansion.

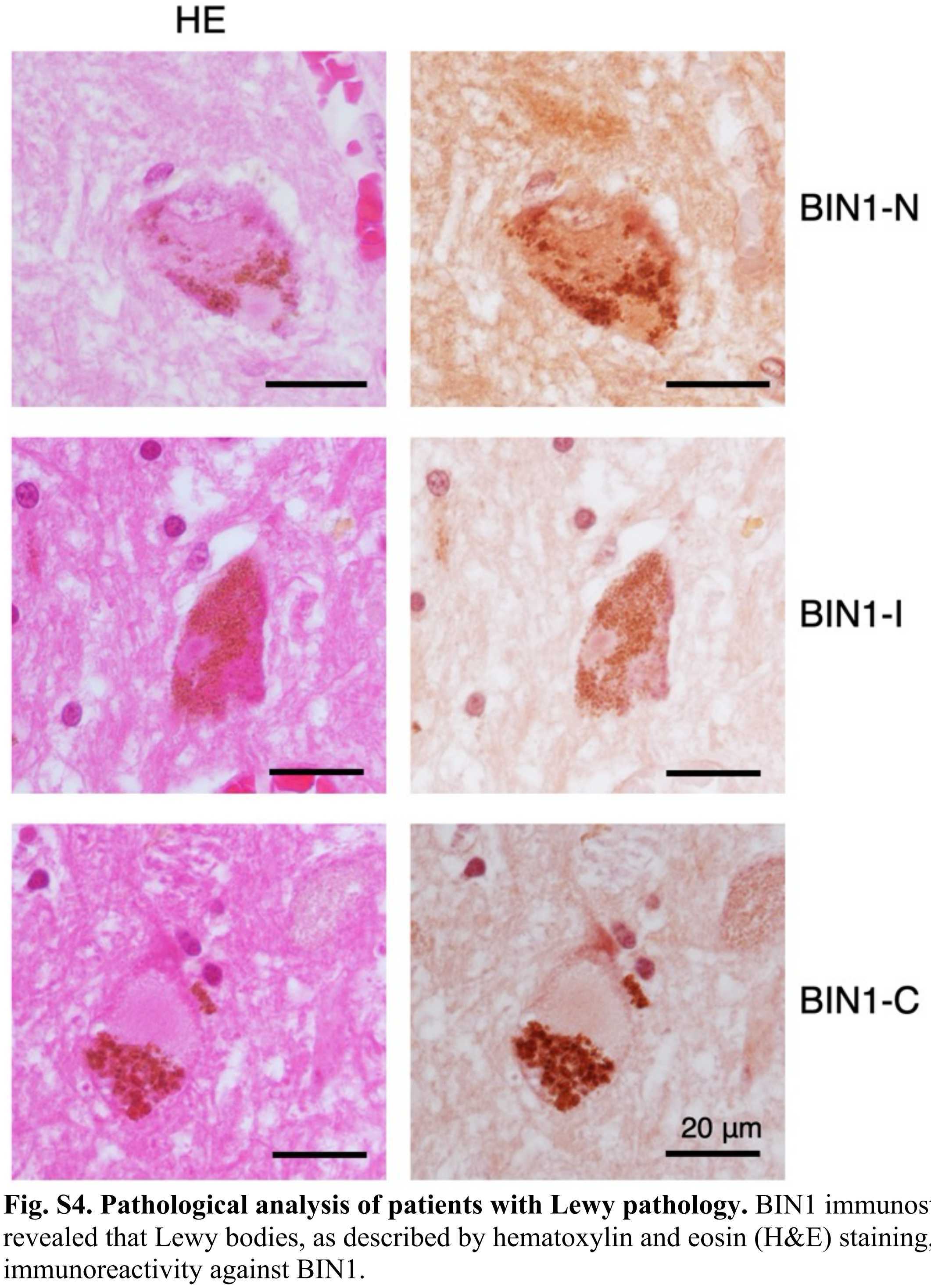


**Fig. S4. Pathological analysis of patients with Lewy pathology.** BIN1 immunostaining revealed that Lewy bodies, as described by hematoxylin and eosin (H&E) staining, show no immunoreactivity against BIN1.

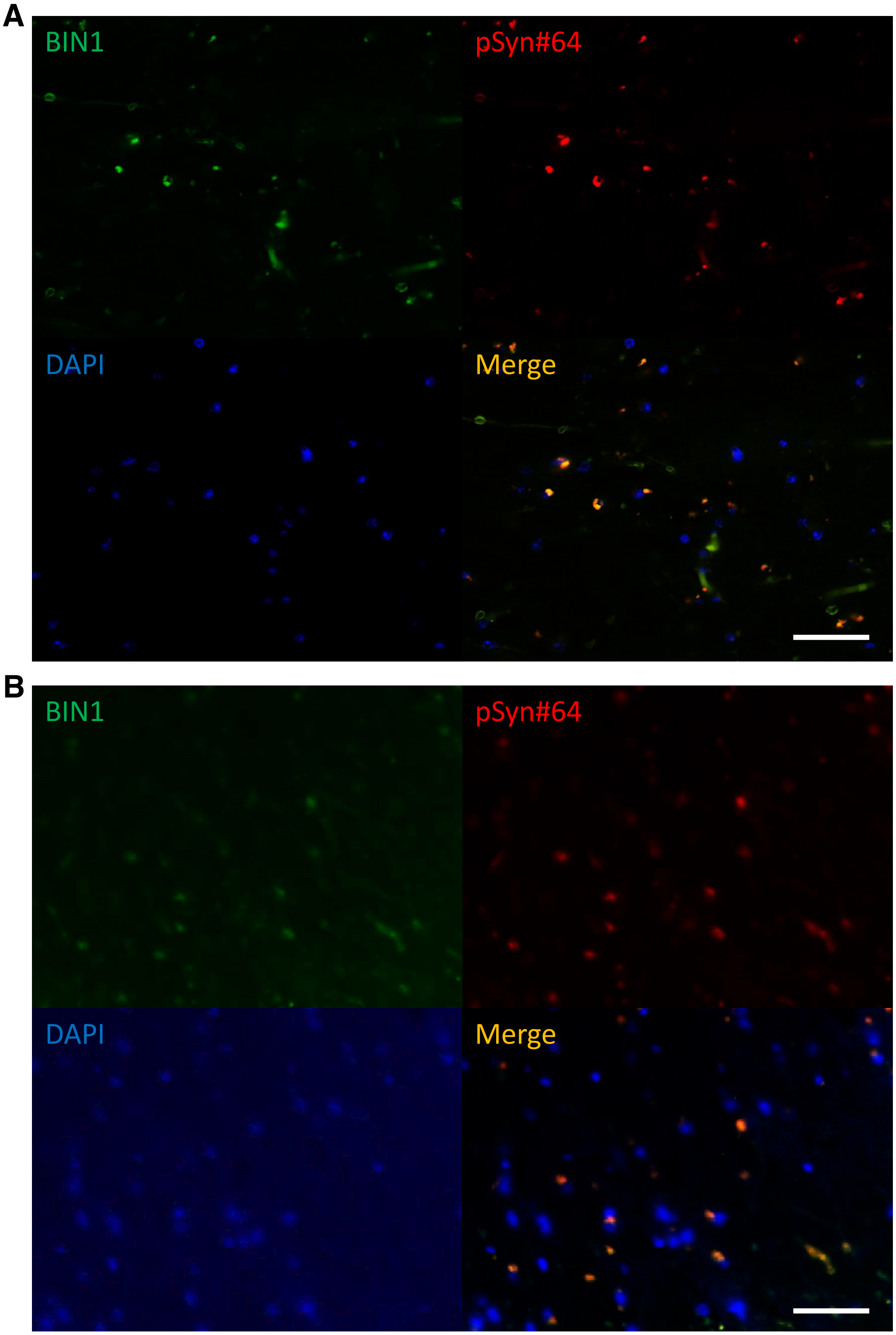


**Fig. S5. Immunofluorescent Analysis of MSA. A.** In MSA (R+) cases, some of the glial cytoplasmic inclusions (GCIs) had immunoreactivity against BIN1 but not pSyn#64. **B,** MSA (R-) showed that almost all GCIs have immunoreactivity against BIN1 and pSyn#64.

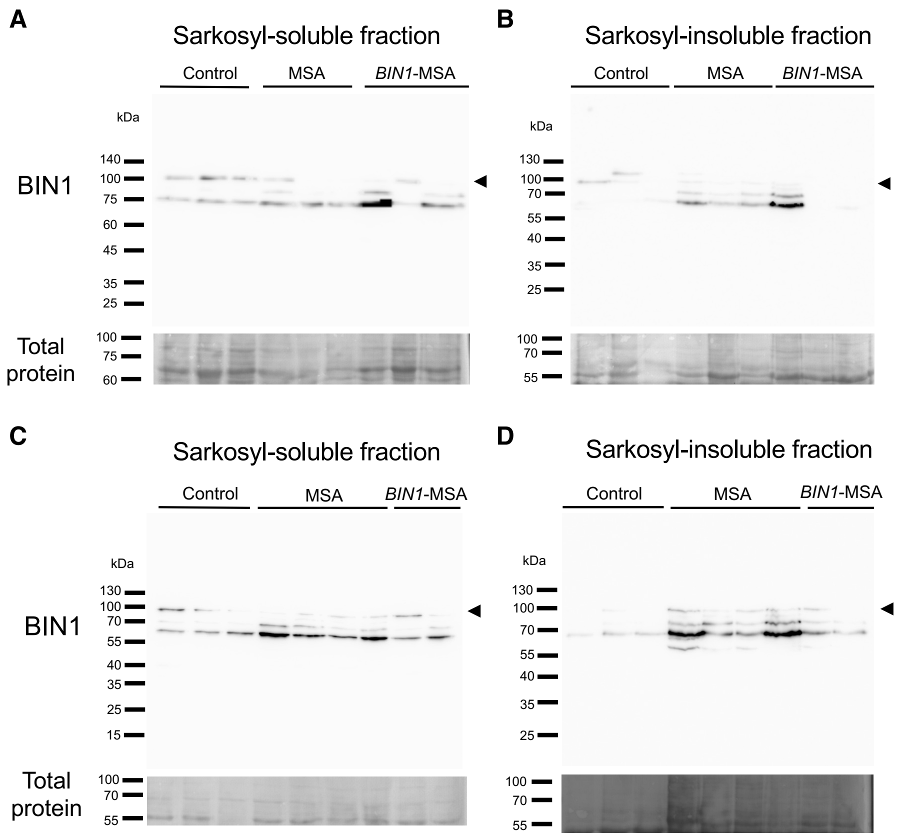


**Fig. S6. Immunoblot analysis of BIN1 in MSA brains (Related to Figure 4).** Immunoblots of BIN1 in the frontal cortex from control subjects (n = 6), individuals with MSA (R-) (n = 7), and MSA (R+) (n = 5). **A, C,** sarkosyl-soluble fraction. **B, D,** sarkosyl-insoluble fraction. The arrowhead indicates the bands corresponding to isoform 1 of BI

**Table S1. Frequency of AGG Repeat Expansion in Patients with MSA.**

| Group | Repeat number | | Frequency | OR<br>95% CI | *p*-value |
|---|---|---|---|---|---|
| | ≥ 80 | < 80 | | | |
| Brain Control | 0 | 65 | 0 | | |
| Pathological MSA | 9 | 58 | 0.134 | Infinity<br>[2.1, Infinity] | 0.003 |
| Blood Control | 14 | 560 | 0.024 | | |
| Clinical MSA | 10 | 214 | 0.045 | 1.87<br>[0.8, 4.5] | 0.16 |

*P*-values are calculated using the Fisher's exact test.
OR: Odds ratio
CI: Confidence interval

**Table S2. Summary of patients with MSA whose brain tissues were histopathologically examined in this study (Related to Figures 4 and 5).**

| Case | Sex | Age at onset | Age at death | Clinical Diagnosis | Repeat number of *BIN1* | Cerebellar myelin pallor | Cerebellar infarct |
|---|---|---|---|---|---|---|---|
| 1 | F | 50s | 60s | MSA-P | 229/9 | + | - |
| 2 | M | 60s | 70s | MSA-C | 243/30 | + | + |
| 3 | M | 60s | 70s | MSA-P | 202/19 | + | - |
| 4 | F | 50s | 70s | MSA-C | 88/11 | + | - |
| 5 | F | 60s | 70s | MSA-C | 12/12 | - | - |
| 6 | M | 80s | 80s | MSA-P | 12/12 | - | - |
| 7 | F | 50s | 60s | MSA-C | 10/10 | - | - |
| 8 | M | 70s | 70s | MSA-C | 11/7 | + | + |
| 9 | M | 60s | 70s | MSA-C | 12/12 | - | - |

F, female, M, male

**Table S3. Summary of patients with Lewy pathology**

| Case | Sex | Age at onset | Age at death | Clinical Diagnosis | Braak PD stage |
|---|---|---|---|---|---|
| 1 | F | NA | 80s | ILBD | 1 |
| 2 | M | 50s | 60s | PD | 3 |
| 3 | M | 60s | 70s | PD | 3 |
| 4 | M | 30s | 40s | PD | 3 |
| 5 | M | 60s | 70s | PD | 3 |
| 6 | M | 60s | 70s | PD | 5 |
| 7 | M | 50s | 70s | PD | 5 |
| 8 | F | 60s | 80s | PDD | 5 |
| 9 | M | 50s | 60s | PDD | 6 |
| 10 | F | 70s | 80s | PDD | 6 |

NA, not available; ILBD, incidental Lewy body disease; PD, Parkinson disease; F, female; M, male

### Table S4. Primer sequences and PCR conditions.

| | Primers and Reagents (Final concentration) | PCR conditions |
|---|---|---|
| RP-PCR | BIN1-RP1: 5'-CAACCTGAAGTCTTCCCAGCCCCCAGCCAG-3' (180 nM) | 95 °C, 5 min |
| | BIN1-RP2: 5'-CCGGGAGCTGCATGTGTCAGAGG AGGAGGAGGAGGAGG-3' (9 nM) | (95 °C, 30 s; 98 °C, 10 s; 62 °C [-0.3 °C/cycle], 30 s; 72 °C, 2 min) x 50 cycle |
| | BIN1-RP3: 5'-FAM-CCGGGAGCTGCATGTGTCAGAGG-3' (180 nM) | 72 °C, 10 min |
| | 2x GC Buffer II (1x) | |
| | dNTP (200 μM) | |
| | 7-deaza-GTP (200 μM) | |
| | LA taq (0.05 U/μL) | |
| Flanking PCR | BIN1-Fw: 5'-CAACCTGAAGTCTTCCCAGCCCCCAGCCAG-3' (0.5 μM) | 98 °C, 1 min |
| | BIN1-Rv: 5'-FAM-CCTGTGTTTCCTGGCTAAGG-3' (0.5 μM) | (95 °C, 30 s; 61 °C, 30 s; 72 °C, 1 min) x 40 cycle |
| | 2x GC Buffer II (1x) | 72 °C, 10 min |
| | dNTP (200 μM) | |
| | 7-deaza-GTP (200 μM) | |
| | LA taq (0.05 U/μL) | |

RP, repeat-primed; FAM, fluorescein

**Table S5. List of primary and secondary antibodies (related to Figures 3–6).**

IHC, Immunohistochemistry; IB, Immunoblot; IEM, Immuno-electron microscopy analysis

| **Antibody** | **Manufacturer** | **Catalogue No.** | **Host** | **Dilution** |
|---|---|---|---|---|
| BIN1-C | Atlas Antibodies | HPA003894 | rabbit | IHC: 400, IB: 1000 |
| BIN1-I | Abcam | EPR13463 | rabbit | 400 |
| BIN1-N | ThermoFisher Scientific | PA5-34687 | rabbit | IHC: 400, IEM: 500 |
| Olig2 | Merck-Millipore | MABN50 | mouse | 100 |
| Phosphorylated α-synuclein (pSyn) | Wako Chemical | pSyn#64 | mouse | 10,000 |
| Alexa Fluorn 488 anti-rabbit IgG | Thermo Fisher Scientific | A11034 | goat | 1,000 |
| Alexa Fluorn 568 anti-mouse IgG | Thermo Fisher Scientific | A11004 | goat | 1,000 |
| Rabbit IgG horseradish peroxidase-conjugated antibody | RD systems | HAF008 | goat | 5000 |
| 12 nm Colloidal Gold anti-rabbit IgG | Jackson ImmunoResearch | 711-205-152 | donkey | 40 |
